# Antiseizure Medication Administration Gaps Across the ICU-to-Floor Transfer: A Matched Within-Patient Comparison

**DOI:** 10.64898/2026.08.26.26361462

**Authors:** Alon Gorenshtein, Yosef Adiniaev, Amir Srour, Eyal Klang, Oved Daniel

## Abstract

**Objective:** Whether a scheduled antiseizure medication (ASM) continues on schedule across the ICU-to-floor transfer has not been characterized. We quantified ASM administration-gap frequency across this transfer and compared it with gap frequency during matched non-transfer intervals in the same patient and drug.

**Methods:** In this retrospective MIMIC-IV (version 3.1) cohort study, we identified epilepsy and status-epilepticus admissions with an ICU stay followed by floor transfer and a scheduled ASM order active at ICU departure. A gap was defined as an interval exceeding 1.5 times the expected dosing interval between the last ICU dose and first floor dose, or no further dose before discharge, and compared with a matched non-transfer control interval in the same patient and drug (paired McNemar test). A multivariable model evaluated six prespecified clinical predictors; sociodemographic variables were summarized descriptively.

**Results:** Among 2,469 ASM transition-by-drug observations (1,583 admissions, 1,335 patients), an administration gap occurred in 251 (10.2%; 95% CI, 8.7%-11.7%). Gap frequency across the transfer exceeded frequency during matched non-transfer control intervals in the same patient and drug: a paired rate difference of 5.8 percentage points (95% CI, 4.4-7.1; 7.5% vs 1.7%; P = 7.3 × 10^-22^) before the transfer and 6.4 percentage points (95% CI, 4.9-7.9; 8.9% vs 2.5%; P = 1.9 × 10^-23^) after. Gap rates were similar for intravenous-available (9.9%) and oral-only (11.4%) drugs (rate difference, 1.5 percentage points; 95% CI, -1.6 to 4.5; P = .34). None of six prespecified predictors reached significance after correction.

**Significance:** An antiseizure medication administration gap occurred in approximately 1 of every 10 drug-transition observations at the ICU-to-floor transfer, exceeding matched non-transfer gap rates by 5.8 to 6.4 percentage points. This transfer-associated excess, rather than any single medication or patient characteristic, supports a structured medication-continuity check.

**Key Points:**

- Antiseizure medication administration gaps occurred in 10.2% of ICU-to-floor drug transitions (95% CI, 8.7%-11.7%).
- Gap frequency was 5.8-6.4 percentage points higher across the transfer than during matched non-transfer control intervals.
- No prespecified clinical predictor or IV-availability class identified a clearly higher-risk subgroup.
- Gap rates were similar for intravenous-available and oral-only medications (9.9% vs 11.4%; difference, 1.5 points; 95% CI, -1.6 to 4.5).
- A structured medication-continuity check at the ICU-to-floor transfer may reduce this administration gap.

## Introduction

Epilepsy affects an estimated 1.2% of the United States population, and seizures and their complications account for a substantial share of acute hospital and intensive care unit (ICU) admissions each year.^1^ Maintaining scheduled ASM therapy is an important component of inpatient seizure management, and medication interruption is clinically concerning because inadequate adherence has been associated with seizure-related morbidity,^2^ though a recent prospective study found occasional missed doses were not independently associated with acute seizure risk in a small cohort with drug-resistant epilepsy.^3^ Patients with epilepsy who require ICU-level care are transferred out of the ICU as their acute illness resolves, a recognized point of vulnerability for medication errors more broadly.^4^

Discontinuation of chronic medications after critical illness has been documented for statins, levothyroxine, and inhaled respiratory therapies using outpatient pharmacy-refill data spanning hospitalization.^5^ Continuation of newly initiated opioid therapy across the ICU-to-floor transition itself has been examined by manual chart review in a single center.^6^ Nonoptimal anticonvulsant medication management during the transition into the hospital has similarly been described in a pediatric case series.^7^ A separate single-center chart review examined antiepileptic drug management strategies during nil-per-os episodes in hospitalized patients with epilepsy.^8^ Whether a scheduled antiseizure medication continues on schedule at ICU transfer, at a scale beyond single-center chart review and using the timestamped electronic medication administration record rather than order-level documentation, has not been characterized.

We conducted a retrospective cohort study using the Medical Information Mart for Intensive Care-IV (MIMIC-IV) database to quantify the frequency of an antiseizure medication administration gap across the ICU-to-floor transition in patients with an epilepsy or status epilepticus diagnosis, and, using a matched comparison against non-transfer control intervals in the same patient and drug, to test whether gap frequency was elevated specifically during the transfer-crossing interval. We also examined the drug-class and clinical characteristics associated with the gap.

## Methods

### Study Design and Setting

This retrospective cohort study used the Medical Information Mart for Intensive Care-IV (MIMIC-IV), version 3.1, a publicly available, deidentified electronic health record database of adult patients admitted to an academic medical center in Boston, Massachusetts, between 2008 and 2022.^9^ The study did not test a causal hypothesis about downstream clinical outcomes; it quantified a care-transition process itself. The study is reported according to the Strengthening the Reporting of Observational Studies in Epidemiology (STROBE) guideline for cohort studies.^10^

### Cohort Identification

Admissions with a diagnosis of epilepsy or recurrent seizures were identified from the diagnoses_icd table using International Classification of Diseases, Ninth Revision (ICD-9) codes beginning with 345 or Tenth Revision (ICD-10) codes beginning with G40, in any diagnosis position. A status epilepticus subgroup was identified as admissions coded with ICD-9 code 345.3 or an ICD-10 G40 code whose descriptor specified status epilepticus, a modifier on the G40 subcode.

Admissions were retained if the patient had at least one ICU stay (icustays table) followed by a transfer to a non-critical-care inpatient ward, identified from the transfers table as the record whose start time exactly matched the ICU stay’s end time and whose care unit was not an ICU, coronary care unit, emergency department, or discharge disposition. Step-down and intermediate-level units were classified as floor destinations, consistent with their de-escalation from critical-care-level monitoring.

A hospitalization was eligible for the primary analysis if a scheduled ASM order was active in the pharmacy record at the exact instant of ICU departure. Active antiseizure medications were levetiracetam, phenytoin (including the prodrug fosphenytoin, treated as pharmacologically equivalent to phenytoin), valproate (including divalproex formulations), lacosamide, phenobarbital, oxcarbazepine, lamotrigine, topiramate, zonisamide, and carbamazepine. Orders with an as-needed (PRN) or single-dose (ONCE) frequency were excluded, as were phenobarbital orders identifiable as an alcohol-withdrawal taper protocol. The expected dosing interval for each order was derived from the pharmacy record’s doses-per-24-hours field.

### Data Sources and Variables

Medication administration times were obtained from the electronic medication administration record (eMAR), restricted to records coded as Administered, Delayed Administered, or Administered Bolus from IV Drip. Administration records were matched to a hospitalization by both subject and hospital admission identifiers.

Covariates were age, sex, race and ethnicity, insurance type, primary language, and marital status (patients and admissions tables); ICU length of stay (icustays); a status epilepticus diagnosis flag (defined above); the count of distinct ASM orders active at ICU departure (polytherapy count); and whether the medication had an intravenous formulation available in this dataset (levetiracetam, phenytoin, valproate, phenobarbital, and lacosamide) or was oral-only (lamotrigine, topiramate, zonisamide, oxcarbazepine, and carbamazepine).

### Outcomes

The primary outcome was an administration gap: for each active ASM order at each qualifying ICU-to-floor transition, the interval between the last ICU administration before transfer and the first floor administration after it, flagged as a gap when that interval exceeded 1.5 times the medication’s expected dosing interval, or when no further administration was confirmed before hospital discharge. The 1.5-times threshold was the primary, prespecified definition; thresholds of 2.0 and 3.0 times the expected interval were examined in sensitivity analyses.

When no further administration was confirmed before discharge, the observation was excluded as not evaluable, rather than counted as a gap, if the next dose’s due time (the last confirmed ICU dose plus the expected dosing interval) fell on or after hospital discharge. This criterion distinguishes a hospitalization that ended before the next dose was due from a true non-resumption gap (Results, Figure 1).

**Figure 1.**
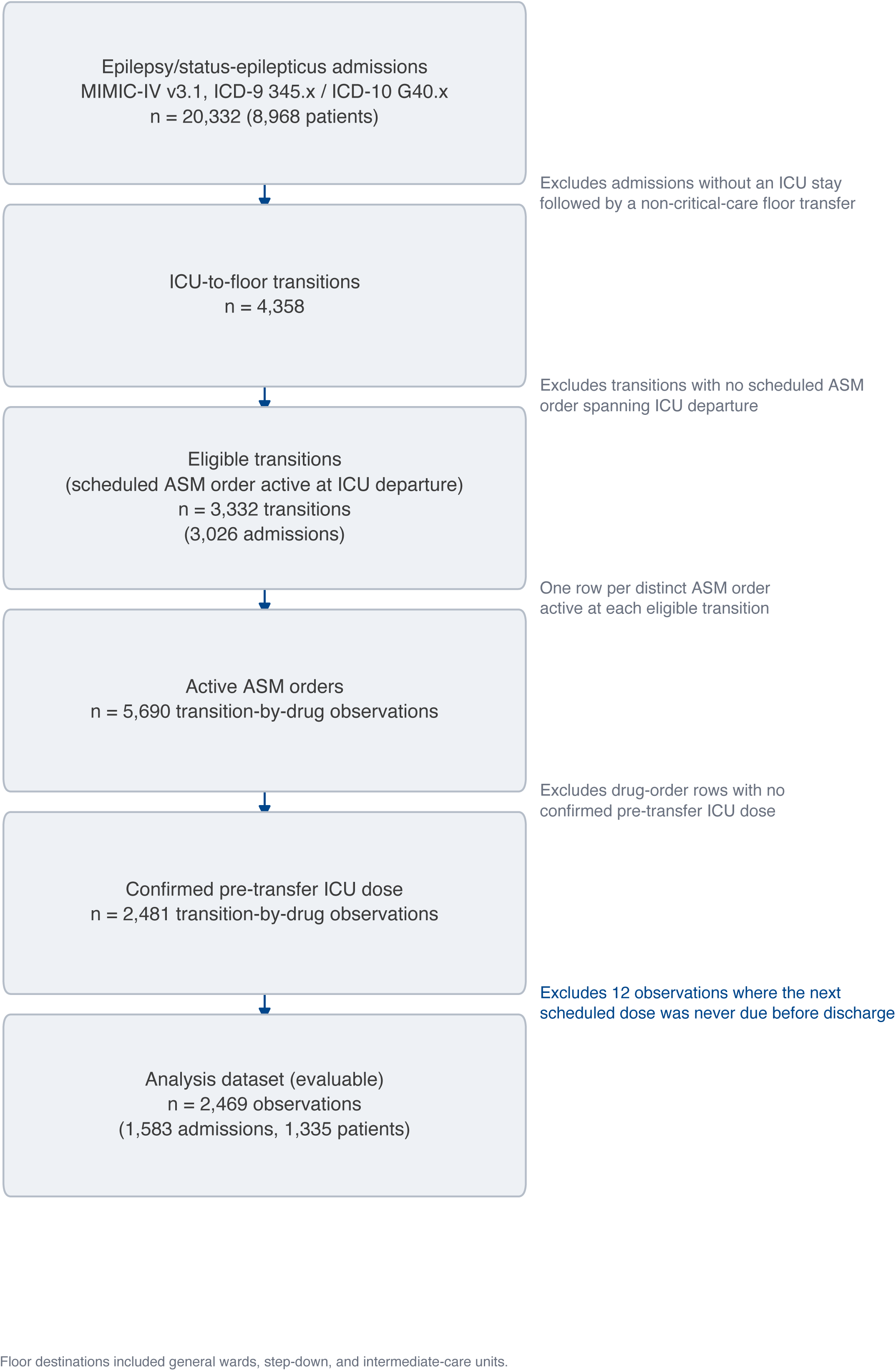
Cohort flow from epilepsy or status-epilepticus admission identification (MIMIC-IV v3.1) through the analysis dataset, with the exclusion reason and count at each step: 20,332 epilepsy or status-epilepticus admissions (8,968 patients); 4,358 ICU-to-floor transitions (3,899 admissions); 3,332 eligible transitions (3,026 admissions) with a scheduled antiseizure medication (ASM) order active at ICU departure; 5,690 active ASM-order (transition-by-drug) observations; 2,481 observations with a confirmed pre-transfer ICU dose; and 2,469 evaluable observations in the analysis dataset (1,583 admissions, 1,335 patients), after excluding 12 observations for which the next scheduled dose was never due before discharge.

A secondary outcome was non-resumption before discharge, defined as an evaluable administration gap in which no further dose of the medication was confirmed before the hospitalization ended.

For each gap, a co-occurring administration of a different antiseizure medication within 24 hours after ICU departure was flagged descriptively as a same-day medication change, an upper bound on apparent medication switching (Study Limitations).

### Matched Non-Transfer Control Intervals

For each evaluable transfer observation, a non-transfer control interval in the same patient and drug was identified, when constructible: a pre-transfer control interval (the second-to-last-to-last ICU-charted dose of the same drug) and a post-transfer control interval (the first-to-second floor-charted dose of the same drug), each flagged as a gap using the identical 1.5-times threshold and compared with the transfer interval by an exact McNemar test on the discordant pairs (eMethods). The expected dosing interval used across the transfer interval and both control intervals was, in every case, the interval derived from the medication order active at the moment of ICU departure, held fixed for a given transition-by-drug observation; a sensitivity check confirmed this rarely differed from the order active during the post-transfer window and gave a similar result in the schedule-unchanged subset (eMethods, eTable 3). The absolute rate difference between the transfer-crossing gap rate and the matched control gap rate was estimated with an admission-cluster bootstrap 95% confidence interval, since an admission or patient could contribute more than one observation to this comparison; a patient-cluster bootstrap served as a sensitivity check (eMethods, eTable 3). These matched-comparator analyses are restricted to the subset of the 2,469 analysis-dataset observations with a constructible control interval, a subset not randomly drawn from the analysis dataset (Results, eTable 3).

### Delay-Duration Distribution

Among administration gaps, the primary summary was the excess delay beyond the expected dosing interval: the raw interdose interval (last confirmed ICU dose to first confirmed floor dose, or discharge if not resumed) minus the expected dosing interval, reported as median, interquartile range, 90th percentile, and maximum, with the raw interdose interval itself reported alongside as a secondary reference. Both summaries were computed separately for resumed and non-resumed gaps (eMethods).

### Statistical Analysis

The primary gap rate, the intravenous-availability comparison, and the matched-comparator rate difference (Matched Non-Transfer Control Intervals) each used an admission-cluster bootstrap 95% confidence interval (5,000 resamples, clustered by hospital admission identifier; eMethods). Wilson score 95% confidence intervals were used for all secondary and exploratory summaries: the sensitivity analyses, the Table 1 stratified rates, the by-drug and dosing-interval-class breakdowns, and the descriptive demographic analysis. Intravenous-availability class and gap occurrence were also compared with a chi-square test of independence (effect size, Cramer V).

**Table 1.** Administration gap rate by clinical and treatment characteristic. Descriptive only; no adjusted or unadjusted P values are reported here, since the six-predictor multivariable model (Multivariable Analysis) is the primary adjusted analysis. The Overall, IV formulation available, and Oral-only rows’ 95% CIs are admission-cluster bootstrap intervals (the Overall row matching the Abstract’s primary estimate; Administration Route); all other rows use a Wilson score interval, consistent with this manuscript’s convention for descriptive/secondary summaries

| Characteristic | n | Gap, n (%) | 95% CI |
| --- | --- | --- | --- |
| Overall | 2,469 | 251 (10.2%) | 8.7%-11.7%<sup>1</sup> |
| Status epilepticus | 585 | 63 (10.8%) | 8.5%-13.5% |
| No status epilepticus | 1,884 | 188 (10.0%) | 8.7%-11.4% |
| Polytherapy | 1,303 | 136 (10.4%) | 8.9%-12.2% |
| Monotherapy | 1,166 | 115 (9.9%) | 8.3%-11.7% |
| IV formulation available | 1,985 | 196 (9.9%) | 8.3%-11.4% |
| Oral-only | 484 | 55 (11.4%) | 8.5%-14.4% |
| Dosed every 6 hours | 42 | 7 (16.7%) | 8.3%-30.6% |
| Dosed every 8 hours | 201 | 56 (27.9%) | 22.1%-34.4% |
| Dosed every 12 hours | 1,844 | 158 (8.6%) | 7.4%-9.9% |
| Dosed every 24 hours | 382 | 30 (7.9%) | 5.6%-11.0% |

<sup>1</sup>Admission-cluster bootstrap 95% CI (5,000 resamples); IV formulation available and Oral-only rows also use the admission-cluster bootstrap (Administration Route); all other rows use a Wilson score 95% CI.

A multivariable logistic regression model (continuous predictors standardized before estimation; odds ratios and 95% confidence intervals from the same 5,000 admission-cluster bootstrap resamples) evaluated six prespecified predictors: drug class, ICU length of stay, status epilepticus, polytherapy count, age, and sex. Two-sided bootstrap P values were adjusted with the Benjamini-Hochberg false discovery rate procedure. Race and ethnicity, insurance type, primary language, and marital status were examined descriptively, with an omnibus chi-square test and Wilson score 95% confidence intervals for each category, rather than as model covariates, because of an insufficient events-per-variable ratio (eMethods).

Four additional analyses accompanied the primary analysis (eMethods): a threshold sensitivity analysis at 2.0 and 3.0 times the expected dosing interval; a one-transition-per-admission analysis; a switch-excluded analysis restricted to observations without a same-day medication-change flag (Outcomes), applied post hoc rather than as a cohort-eligibility criterion; and an admission-cluster-adjusted rate difference (oral-only minus intravenous-available; Administration Route).

All analyses were performed in Python 3.9 with pandas 2.x, numpy, scikit-learn, scipy, and statsmodels 0.14. Statistical significance threshold was *P* < .05.

### Ethics

This study used MIMIC-IV, a publicly available, deidentified database. The data use agreement, together with completion of required human-subjects research training by the investigators, substitutes for site-specific institutional review board approval for secondary analyses; individual patient consent was waived by the data provider because all records were deidentified before release. No separate institutional review board approval was required for this secondary analysis of a public, deidentified dataset.

### Data Availability

MIMIC-IV is publicly available to credentialed users through PhysioNet (physionet.org). Analysis code is available at https://github.com/Alon-Gorenshtein/ASM_Continuity.

### Funding

None.

### Author Contributions

Conception and study design: A.G. Data acquisition and analysis: A.G., Y.A., A.S. Drafting the manuscript: A.G. Critical revision for important intellectual content: all authors. Supervision: E.K., O.D.

### Ethical Publication Statement

We confirm that we have read the Journal’s position on issues involved in ethical publication and affirm that this report is consistent with those guidelines.

## Results

### Cohort

Among 20,332 hospital admissions with a diagnosis of epilepsy or recurrent seizures in MIMIC-IV, version 3.1 (8,968 unique patients), 973 admissions (828 patients) carried a status epilepticus diagnosis. A total of 4,358 ICU-to-floor transitions (3,899 admissions) were identified. Of these transitions, 3,332 (3,026 admissions) had a scheduled ASM order active at the exact instant of ICU departure and were eligible for the primary analysis, generating 5,690 transition-by-drug observations (one row per ASM order active at each eligible transition). Restricting to transition-by-drug observations with a medication administration confirmed in the ICU before transfer left 2,481 observations. The 3,209 excluded observations reflect an active ASM order with no administration confirmed in the eMAR before transfer (for example, an order not yet due for its first dose, or an administration recorded under an uncaptured drug name), not a documented gap; the analysis population is accordingly scheduled ASMs with confirmed pre-transfer administration, not all ASM orders active at transfer. Of the 2,481 observations, 12 (0.5%) were excluded as not evaluable because the next scheduled dose was never due before hospital discharge (Methods), leaving 2,469 observations in the analysis dataset (1,583 admissions, 1,335 patients) (Figure 1). Cohort and treatment characteristics, and the administration gap rate within each, are summarized in Table 1.

Levetiracetam was the most frequently observed active medication (1,274 observations, 51.6%), followed by lacosamide (256, 10.4%), phenytoin (229, 9.3%), lamotrigine (190, 7.7%), valproate (165, 6.7%), oxcarbazepine (100, 4.1%), topiramate (67, 2.7%), zonisamide (65, 2.6%), carbamazepine (62, 2.5%), and phenobarbital (61, 2.5%). Median (IQR) age was 59 (49-69) years, 1,254 observations (50.8%) were in male patients, and the median (IQR) concurrent ASM count was 2 (1-2).

### Matched Non-Transfer Comparator

#### The administration gap was substantially more frequent during the ICU-to-floor transfer interval than during background non-transfer intervals in the same patient and drug

Among 2,247 observations with a constructible pre-transfer control interval (Methods), a gap occurred across the transfer but not during the matched control interval in 163 observations, and a gap occurred during the control interval but not across the transfer in 33 observations (6 observations had a gap in both intervals, 2,045 in neither); the transfer-crossing gap rate (7.5%) exceeded the pre-transfer control gap rate (1.7%; exact McNemar test, P = 7.26 × 10^-22^). The paired rate difference was 5.8 percentage points (95% CI, 4.4-7.1; admission-cluster bootstrap). Among 2,356 observations with a constructible post-transfer control interval (Methods), the pattern was similar: a gap occurred across the transfer but not during the control interval in 197 observations, and during the control interval but not across the transfer in 46 observations (13 in both, 2,100 in neither); the transfer-crossing gap rate (8.9%) exceeded the post-transfer control gap rate (2.5%; P = 1.92 × 10^-23^). The paired rate difference was 6.4 percentage points (95% CI, 4.9-7.9; admission-cluster bootstrap). Both comparisons paired the transfer interval against a control interval from the same patient and drug, showing an excess gap rate tied to the transfer-crossing interval itself (Figure 2a; eTable 3).

**Figure 2.**
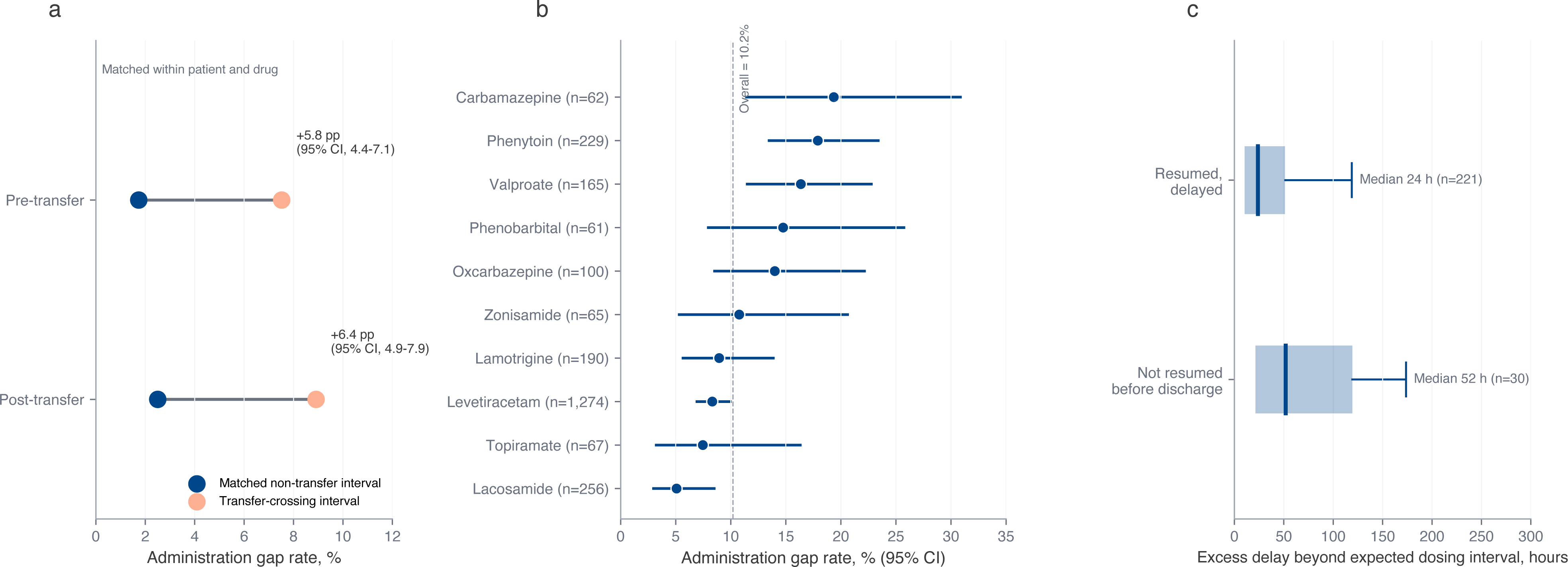
(a) Antiseizure medication administration gap rate across the ICU-to-floor transfer versus the matched non-transfer control interval, for the pre-transfer and post-transfer control comparisons, each a paired comparison in the same patient and drug (exact McNemar test); the annotation for each pair gives the paired rate difference in percentage points, with its admission-cluster bootstrap 95% confidence interval. (b) Gap rate by antiseizure medication, with 95% Wilson confidence intervals, sorted from lowest to highest; the dashed line marks the overall cohort gap rate (10.2%). (c) Distribution of excess duration beyond the expected dosing interval among administration gaps, comparing gaps resumed before discharge with gaps never resumed before discharge (median, interquartile-range bar, and 90th percentile). The six-predictor multivariable logistic-regression model (Results, Multivariable Analysis) is reported in-text and is not separately paneled in this figure; the descriptive demographic breakdown by race, language, insurance, and marital status is reported in eTable 1 of the Supplement.

These comparisons are restricted to the subsets of the 2,469 analysis-dataset observations with a constructible control interval: 2,247 (91.0%) for the pre-transfer comparison and 2,356 (95.4%) for the post-transfer comparison. Observations without a constructible control interval had a substantially higher gap rate than those with one: 36.9% (95% CI, 30.9%-43.5%) versus 7.5% (95% CI, 6.5%-8.7%) for the pre-transfer comparison, and 36.3% (95% CI, 28.0%-45.5%) versus 8.9% (95% CI, 7.8%-10.1%) for the post-transfer comparison. For the post-transfer comparison, this pattern has an identifiable mechanism: a post-transfer control interval requires two floor-charted doses of the same drug, so a gap never resumed before discharge, which by definition has no floor-charted dose, and some early-discontinuation observations cannot generate one; the post-transfer comparator’s evaluable subset is therefore enriched for observations with resumed therapy; the matched estimates above apply to this control-evaluable subset rather than to the full analysis dataset. A similar mechanism may partly explain the pre-transfer pattern (a drug recently initiated in the ICU, with only a single ICU-charted dose before transfer, cannot generate a pre-transfer control interval), though this candidate explanation has not been directly verified against order-start timestamps (eTable 3).

### Primary Outcome

#### A scheduled antiseizure medication was not re-administered on schedule across the ICU-to-floor transfer in approximately 1 of every 10 drug-transition observations

An administration gap (Methods) occurred in 251 of 2,469 observations (10.2%; 95% CI, 8.7%-11.7%; admission-cluster bootstrap). Among these 251 gaps, 30 (12.0% of gaps, 1.2% of the cohort) reflected no further dose of the medication confirmed before discharge; the remaining 221 (88.0% of gaps, 8.9% of the cohort) reflected a delayed, not absent, resumption.

### Delay to Resumption

Among the 251 administration gaps, the excess delay beyond the expected dosing interval had a median (IQR) of 24.2 (10.5-58.7) hours (90th percentile, 131.0 hours; maximum, 710.5 hours), corresponding to a raw interdose interval (Methods) of 37.2 (22.6-71.6) hours (90th percentile, 143.0 hours; maximum, 722.5 hours). Gaps resumed before discharge (n = 221) had a shorter and less variable excess delay (median [IQR], 23.8 [10.4-51.2] hours; 90th percentile, 118.8 hours) than gaps never resumed before discharge (n = 30; median [IQR], 51.9 [21.4-119.3] hours; 90th percentile, 173.7 hours) (Figure 2c; eTable 4).

### Multivariable Analysis

In the multivariable logistic regression model of the six prespecified predictors (Methods), none reached significance after Benjamini-Hochberg correction. Odds ratios (per SD for continuous predictors) were: age, 1.14 (95% CI, 0.96-1.37; P = .13; adjusted P = .40); ICU length of stay, 0.81 (95% CI, 0.57-1.02; P = .08; adjusted P = .40); oral-only drug class, 1.07 (95% CI, 0.93-1.22; P = .31; adjusted P = .62); polytherapy count, 1.03 (95% CI, 0.86-1.22; P = .73; adjusted P = .73); status epilepticus, 1.08 (95% CI, 0.89-1.30; P = .41; adjusted P = .62); and male sex, 0.95 (95% CI, 0.80-1.12; P = .51; adjusted P = .62). Race and ethnicity, insurance type, primary language, and marital status were examined descriptively rather than as covariates (Methods); gap rates by category are summarized in eTable 1.

### Administration Route

The gap rate was 9.9% (95% CI, 8.3%-11.4%; admission-cluster bootstrap) for the 1,985 observations with an intravenous formulation available and 11.4% (95% CI, 8.5%-14.4%) for the 484 oral-only observations, not statistically significant by chi-square test (Cramer V, 0.018; P = .37). An admission-cluster-adjusted rate difference (oral-only minus intravenous-available) gave a similar result: 1.5 percentage points (95% CI, -1.6 to 4.5; P = .34).

### By-Drug and Dosing-Interval Variation

#### Gap rates varied across the ten antiseizure medications; the most common drug did not drive the headline rate

Gap rates ranged from 5.1% (95% CI, 3.0%-8.5%; n = 256) for lacosamide to 19.4% (95% CI, 11.4%-30.9%; n = 62) for carbamazepine (Figure 2b; eTable 5). Levetiracetam, the most frequent medication (1,274 observations, 51.6% of the cohort), had a gap rate of 8.3% (95% CI, 6.9%-10.0%), below the overall 10.2%; the highest-rate drugs, carbamazepine (19.4%), phenytoin (17.9%), and valproate (16.4%), were each small contributors to the cohort. The remaining drugs fell between these extremes: phenobarbital (14.8%), oxcarbazepine (14.0%), zonisamide (10.8%), lamotrigine (8.9%), and topiramate (7.5%) (eTable 5).

#### The gap rate varied by expected dosing interval; medications dosed every 8 hours showed a higher rate than the two most common interval classes

The gap rate was 27.9% (95% CI, 22.1%-34.4%; n = 201) for medications dosed every 8 hours, more than three times 8.6% (95% CI, 7.4%-9.9%; n = 1,844) for every 12 hours and 7.9% (95% CI, 5.6%-11.0%; n = 382) for every 24 hours. Medications dosed every 6 hours (n = 42, the smallest interval class) had a gap rate of 16.7% (95% CI, 8.3%-30.6%). This pattern is descriptive, an association rather than a causal effect of dosing frequency (eTable 4).

### Sensitivity Analyses

The gap rate at the primary 1.5-times threshold was 10.2% (95% CI, 9.0%-11.4%; Wilson score interval), consistent with the admission-cluster bootstrap estimate reported above. As expected by construction, the rate decreased as the threshold defining a gap was loosened: 7.1% (95% CI, 6.1%-8.2%) at 2.0 times the expected dosing interval and 5.5% (95% CI, 4.7%-6.5%) at 3.0 times (eTable 2). Restricting the cohort to one transition per hospital admission gave a gap rate of 11.1% (95% CI, 9.7%-12.8%; n = 1,583 admissions). Excluding the 1,373 observations flagged with a same-day medication change (Methods) and restricting to the remaining 1,096 observations gave a gap rate of 10.8% (95% CI, 9.1%-12.7%). This flag, present for 1,373 of 2,469 observations (55.6%), reflects polytherapy (52.8% of the cohort) more than genuine single-drug substitution. The matched-comparator excess also persisted at the 2.0-times and 3.0-times thresholds, both before and after transfer (all McNemar P < 10^-13^; eTable 3). A ward-only sensitivity analysis (n = 2,250; excludes step-down/intermediate destinations) gave a similar gap rate, 10.8% (95% CI, 9.6%-12.1%), with the matched-comparator excess persisting before and after transfer (eTable 2, eTable 3).

## Discussion

In this retrospective cohort of 2,469 antiseizure medication (ASM) transition-by-drug observations from patients with epilepsy or status epilepticus transferred from the ICU to a general hospital floor, an administration gap occurred in approximately 1 of every 10 observations (10.2%). The central evidence is a matched comparison showing this gap was disproportionately concentrated around the transfer interval: gap frequency across the transfer exceeded gap frequency during non-transfer control intervals from the same patient and drug, both before and after the transfer (Results). Because each comparison held patient and drug constant, the excess gap rate is consistent with a vulnerability specific to the transfer process rather than the patient or drug population.

Consistent with this pattern, none of the six prespecified predictors (Methods) reached significance after correction, and the gap rate was similar for medications with and without an intravenous formulation. A gap concentrated in a particular drug class, a particular illness-severity proxy, or a particular demographic subgroup would point toward a targetable failure point. The absence of such a concentration does not, on its own, identify the transfer process as the point of vulnerability; that identification comes from the matched comparator above.

Discontinuation of chronic medications after critical illness has been documented for statins, levothyroxine, and inhaled respiratory therapies using outpatient pharmacy-refill data spanning hospitalization, which captures resumption after discharge but not events during the stay itself.^5^ Continuity of newly initiated opioid therapy across the same ICU-to-floor transition point has been studied by manual chart review in a single center (n = 112).^6^ The present study applied a comparable framework to antiseizure medications at a larger scale (2,469 transition-by-drug observations) using timestamped administration data rather than manual chart abstraction. A discharge medication-reconciliation audit at a single epilepsy monitoring unit found antiseizure medication errors in 9.9% of admissions, a comparable order of magnitude to the gap rate observed here despite differing methodology and setting.^11^ A separate single-center chart review of antiepileptic drug management during nil-per-os episodes addressed a different scenario, oral-intake restriction, rather than the ICU-to-floor handoff itself.^8^

The matched-comparator finding supports a structured medication-continuity check at the transfer interval itself, built into the ICU discharge or floor-arrival workflow, flagging any ASM order without confirmed administration since the last scheduled interval. This approach would not require changing which medications are prescribed, and established remedial-dosing guidance exists once a gap is identified.^12^ The oral-only ASMs in this cohort showed a numerically higher gap rate than medications with an intravenous option, though not statistically significant; because an oral-only agent has no parenteral bridge if a dose is missed during transfer, this observation may still merit attention in designing such a check, though it does not establish which medications should be prioritized.

### Study Limitations

This study has several limitations. First, MIMIC-IV reflects care at a single academic medical center, so the gap rate reported here may not generalize elsewhere. Second, the eMAR-based gap definition cannot distinguish an unintentional lapse from a clinically appropriate hold. Third, the study did not measure any downstream clinical outcome, such as breakthrough seizure or status epilepticus recurrence, because MIMIC-IV does not reliably capture seizure events as a validated outcome. Fourth, the matched-comparator design does not establish that a transfer-crossing gap carries a different clinical consequence than a similar-duration non-transfer gap. Fifth, a self-controlled comparison cannot rule out residual confounding: control intervals share the same patient, drug, and hospitalization as the transfer interval, but not necessarily the same acuity trajectory. Sixth, cohort identification relied on ICD diagnosis codes, which may misclassify some admissions. Seventh, the same-day medication-change flag could not distinguish a genuine substitution from ordinary concurrent administration in a patient on polytherapy.

### Future Directions

A prospective, multicenter replication would establish whether this excess gap rate generalizes beyond a single center. A study that adjudicates clinical severity at each interval, transfer and matched control, would test whether the excess gap rate persists once acuity trajectory is accounted for. A direct evaluation of an electronic health record-embedded medication-continuity check at ICU discharge would test whether this is actionable. Where a validated seizure-recurrence outcome is available, linking gaps to subsequent breakthrough seizures would address the clinical-consequence question this study could not answer.

### Conclusion

Across the ICU-to-floor transfer, an antiseizure medication administration gap occurred in approximately 1 of every 10 drug-transition observations (10.2%). Gap frequency was higher across the transfer than during matched non-transfer control intervals in the same patient and drug, both before the transfer (7.5% vs 1.7%) and after it (8.9% vs 2.5%), a pattern consistent with a vulnerability specific to the transfer process rather than any single medication or patient characteristic. These findings support directing a structured medication-continuity check to the ICU-to-floor transfer interval.

## Supporting information

Supplementary Material (eMethods and eTables 1-5)

## Data Availability

All data analyzed in the present study are derived from MIMIC-IV (version 3.1), which is publicly available to credentialed users through PhysioNet (https://physionet.org). Access requires completion of the required human-subjects research training and acceptance of the PhysioNet data use agreement. The analysis code is available at https://github.com/Alon-Gorenshtein/ASM_Continuity.

