## Supplementary Material (eMethods and eTables 1-5) for "Antiseizure Medication Administration Gaps Across the ICU-to-Floor Transfer: A Matched Within-Patient Comparison"

This Supplementary Material accompanies the main manuscript. It contains additional statistical detail (eMethods) and five supplementary tables (eTables 1-5) presenting analyses that were summarized or omitted from the main text for length. Race and ethnicity, primary language, insurance type, and marital status are not covariates in the six-predictor multivariable model reported in the main text (Methods); each is instead examined descriptively (eTable 1).

**Contents**

eMethods. Additional statistical detail

eTable 1. Antiseizure medication administration gap rate by race and ethnicity, primary language, insurance type, and marital status

eTable 2. Sensitivity analyses of the primary gap-rate estimate

eTable 3. Matched non-transfer control-interval comparison, full contingency detail

eTable 4. Delay-duration distribution among administration gaps

eTable 5. Antiseizure medication administration gap rate by medication

### eMethods. Additional statistical detail

#### Admission-cluster bootstrap procedure

The primary gap-rate estimate, the admission-cluster-adjusted intravenous-availability rate difference, and the odds ratios and 95% confidence intervals for the six-predictor multivariable logistic regression model were each derived from 5,000 bootstrap resamples of the analysis dataset (n = 2,469 transition-by-drug observations), sampling with replacement and clustered by hospital admission identifier (hadm_id), so that all transition-by-drug observations contributed by the same hospital admission were resampled together rather than independently. This clustering step accounts for a single hospital admission contributing more than one transition-by-drug observation, which occurs when more than one antiseizure medication was active at ICU departure or when an admission included more than one qualifying ICU stay.

For the multivariable model, continuous predictors were standardized before estimation and a logistic regression model with the six prespecified predictors, drug class, ICU length of stay, status epilepticus, polytherapy count, age, and sex, was refit to each cluster-resample. The reported odds ratio for each predictor is the point estimate from the model fit to the full analysis dataset; the 95% confidence interval is the 2.5th and 97.5th percentile of the corresponding bootstrap distribution. A two-sided bootstrap *P* value was calculated for each predictor as twice the smaller of the proportion of bootstrap odds ratios falling above 1 or below 1; the resulting six *P* values were adjusted jointly for multiple comparisons with the Benjamini-Hochberg false discovery rate procedure.

Wilson score 95% confidence intervals, rather than the admission-cluster bootstrap, were used for the sensitivity analyses (eTable 2), the Table 1 stratified rates, the individual-drug and dosing-interval-class breakdowns (eTable 4, eTable 5), and the descriptive demographic analysis (eTable 1), each a secondary or exploratory summary rather than the primary inferential estimate. The transfer-versus-control comparison's rate difference (eTable 3) instead used a dedicated admission-cluster bootstrap 95% confidence interval, described in the next paragraph.

The absolute rate difference between the transfer-crossing gap rate and the matched control gap rate (eTable 3) was derived from 5,000 bootstrap resamples, sampling with replacement and clustered by hospital admission identifier (hadm_id) as the primary method, with a separate bootstrap clustered by patient identifier (subject_id) as a sensitivity check. Each resample was drawn from the subset of observations with a constructible control interval for that comparison (2,247 of 2,469 observations for the pre-transfer comparison, 2,356 of 2,469 for the post-transfer comparison), not from the full analysis dataset, because the rate difference itself is defined only on that subset. The reported rate difference is the point estimate calculated on the full subset; the 95% confidence interval is the 2.5th and 97.5th percentile of the corresponding bootstrap distribution. This bootstrap procedure applies only to the rate-difference confidence interval, the effect-size estimate; the McNemar *P* value for the same comparison (eTable 3) is reported as an exact, two-sided binomial test on the discordant pairs, while the cluster-bootstrap confidence interval, not the McNemar *P* value, provides the primary clustering-aware uncertainty estimate for the absolute rate difference.

#### Matched control-interval design and exact McNemar test

For each evaluable transfer transition-by-drug observation, a non-transfer control interval was identified using the identical gap-threshold rule applied to the primary transfer interval (1.5 times the medication's expected dosing interval). A pre-transfer control interval was constructed from the interval between the second-to-last and the last ICU-charted dose of the same drug, both before the transfer; a post-transfer control interval was constructed from the interval between the first and second floor-charted dose of the same drug, both after the transfer.

A control interval could not be constructed for every observation, for example when the last ICU dose before transfer was also the first dose of that drug charted for the admission, so the pre-transfer and post-transfer comparisons were each restricted to the subset of observations with a constructible control interval of that type (eTable 3). Each transfer interval and its corresponding control interval were paired within the same patient and drug, and each was independently flagged as a gap using the same 1.5-times threshold. The difference in gap frequency between the transfer interval and its control interval was tested with an exact McNemar test, a two-sided binomial test on the discordant pairs (observations with a gap across the transfer but not during the control interval, versus observations with a gap during the control interval but not across the transfer), which does not rely on a large-sample chi-square approximation and is appropriate for the small concordant-gap cell counts observed here (eTable 3).

#### Dosing-interval-schedule sensitivity check

The expected dosing interval used to flag a gap, applied identically to the transfer interval, the pre-transfer control interval, and the post-transfer control interval, was in every case the interval derived from the medication order active at the instant of ICU departure; this single expected interval was held fixed across all three comparisons for a given transition-by-drug observation, rather than re-derived from whichever order happened to be active at the time of each individual interval. Because the post-transfer control interval falls temporally after the ICU-departure order, a later floor-based order for the same drug could in principle specify a different dosing frequency, which would make the ICU-departure-derived expected interval an imprecise yardstick for the post-transfer comparison specifically. A sensitivity check evaluated this directly: of 2,300 post-transfer control observations for which the floor-active order at the time of the post-transfer control interval could be identified and checked against the ICU-departure order, the expected dosing interval differed in 14 (0.6%) and was unchanged in the remaining 2,286 (99.4%). Restricting the post-transfer comparison to this schedule-unchanged subset gave a materially similar result to the full post-transfer comparison (eTable 3): a transfer-crossing gap rate of 8.7% versus a control gap rate of 2.4% (exact McNemar test, *P* = 8.27 × 10^-23^).

#### Control-evaluable-subset selection check

Because a control interval could not be constructed for every analysis-dataset observation, the pre-transfer and post-transfer comparisons are each restricted to a subset of the 2,469-observation analysis dataset, and that subset is not a random sample of it. To characterize this restriction, the administration-gap rate among observations with a constructible control interval, the subset entering the eTable 3 comparator analysis, was compared with the gap rate among observations without one, separately for the pre-transfer and post-transfer control types, each with a Wilson score 95% confidence interval (eTable 3). For the post-transfer comparison, this restriction has an identifiable mechanism: constructing a post-transfer control interval requires two floor-charted doses of the same drug after transfer, so an observation whose gap was never resumed before discharge, which by definition has no floor-charted dose, cannot generate one, nor can some early-discontinuation observations; the post-transfer control-evaluable subset is accordingly enriched for observations with resumed therapy; the transfer-versus-control comparison restricted to it therefore characterizes this control-evaluable subset rather than the full analysis dataset. A related mechanism may partly explain the analogous pre-transfer pattern, a drug recently initiated in the ICU with only a single ICU-charted dose before transfer cannot generate a pre-transfer control interval, though this candidate explanation has not been directly verified against order-start timestamps.

#### Delay-duration summary statistics

Among the 251 observations flagged as an administration gap, the primary summary was the excess delay beyond the expected dosing interval, calculated for each gap as the raw interdose interval, the interval in hours between the last confirmed ICU dose and the first confirmed floor dose, or, when no further dose of that medication was confirmed before discharge, the interval between the last confirmed ICU dose and hospital discharge, minus that observation's own expected dosing interval, and summarized with the median, interquartile range (25th to 75th percentile), 90th percentile, and maximum. The raw interdose interval itself, without the expected-interval subtraction, was reported alongside as a secondary, absolute-elapsed-time reference. Both summaries were computed separately for the 221 gaps in which the medication was resumed before discharge and the 30 gaps in which it was not (eTable 4), because these two groups represent different severities of the same gap category.

#### Descriptive demographic analysis

Race and ethnicity, primary language, insurance type, and marital status were not included as covariates in the six-predictor multivariable logistic regression model (Methods, main text). Fitting these four fields as regression covariates, after expansion into indicator terms, would have required 45 additional terms beyond the six prespecified predictors; with 251 gap events in the analysis dataset, the resulting ratio, approximately 5.6 events per additional indicator term, is below the commonly cited threshold of 10 events per variable for stable maximum-likelihood coefficient estimation, and several of the underlying category levels are themselves too sparsely observed in this cohort to support a stable individual coefficient. Each of the four fields was instead examined descriptively (eTable 1).

Race and ethnicity, which is recorded in MIMIC-IV at a fine-grained subcategory level, was collapsed into five categories for this descriptive analysis: Asian, Black/African American, Hispanic/Latino, White, and Other/Unknown, the last combining all remaining subcategories, including declined and unable-to-obtain responses. Primary language was collapsed into three categories: English, Non-English/Other, and Unknown. Insurance type (Medicaid, Medicare, Other, Private) and marital status (divorced, married, single, widowed) were examined at the categories recorded in MIMIC-IV without further collapsing.

For each of the four fields, an omnibus chi-square test of independence was used to test for an association between the field's categories and gap occurrence, and a Wilson score 95% confidence interval was calculated for the gap rate within each individual category (eTable 1). Category counts do not sum to the full analysis dataset (n = 2,469) for insurance type and marital status, because of missing values for those fields in the underlying MIMIC-IV admission record.

#### Sensitivity analyses

The one-transition-per-admission sensitivity analysis (eTable 2) restricted the analysis dataset to one transition-by-drug observation per hospital admission, retaining the first qualifying transition chronologically when an admission contributed more than one, to assess whether hospitalizations with multiple ICU stays, or with multiple concurrent antiseizure medications each generating its own transition-by-drug observation, affected the primary gap-rate estimate through non-independent observations.

The threshold sensitivity analyses (eTable 2) recalculated the gap flag using the same last-ICU-dose-to-first-floor-dose interval but at administration-interval thresholds of 2.0 and 3.0 times the expected dosing interval, instead of the primary 1.5-times threshold, without altering cohort membership.

The switch-excluded sensitivity analysis (eTable 2) restricted the analysis dataset to the 1,096 observations without a same-day medication-change flag (a co-occurring administration of a different antiseizure medication within 24 hours after ICU departure, Methods, main text), excluding 1,373 observations with that flag, and recalculated the gap rate on the remaining observations.

#### Software

All analyses were performed in Python 3.9 with pandas 2.x, scikit-learn, scipy, and statsmodels 0.14.

### eTable 1. Antiseizure medication administration gap rate by race and ethnicity, primary language, insurance type, and marital status

*This table is descriptive and exploratory, not inferential: no adjusted odds ratios are reported, and none of these four fields is a covariate in the six-predictor multivariable model reported in the main text (Methods, eMethods). For each field, category counts, gap events, gap rate, and a Wilson score 95% confidence interval are shown, along with an omnibus chi-square test of independence for that field. Category counts for insurance type and marital status do not sum to the full analysis dataset (n = 2,469) because of missing values for those fields.*

#### Race and ethnicity

| Category | n | Gap events | Gap rate | 95% CI |
| --- | --- | --- | --- | --- |
| Asian | 53 | 4 | 7.5% | 3.0%, 17.9% |
| Black/African American | 428 | 52 | 12.1% | 9.4%, 15.6% |
| Hispanic/Latino | 92 | 9 | 9.8% | 5.2%, 17.6% |
| White | 1,434 | 151 | 10.5% | 9.0%, 12.2% |
| Other/Unknown | 462 | 35 | 7.6% | 5.5%, 10.4% |

*Omnibus chi-square test of independence: chi-square = 5.86, df = 4, P = 0.210.*

#### Primary language

| Category | n | Gap events | Gap rate | 95% CI |
| --- | --- | --- | --- | --- |
| English | 2,275 | 235 | 10.3% | 9.1%, 11.6% |
| Non-English/Other | 189 | 16 | 8.5% | 5.3%, 13.3% |
| Unknown | 5 | 0 | 0.0% | 0.0%, 43.4% |

*Omnibus chi-square test of independence: chi-square = 1.23, df = 2, P = 0.540.*

#### Insurance type

| Category | n | Gap events | Gap rate | 95% CI |
| --- | --- | --- | --- | --- |
| Medicaid | 529 | 48 | 9.1% | 6.9%, 11.8% |
| Medicare | 1,424 | 154 | 10.8% | 9.3%, 12.5% |
| Private | 455 | 42 | 9.2% | 6.9%, 12.2% |
| Other | 55 | 7 | 12.7% | 6.3%, 24.0% |

*Omnibus chi-square test of independence: chi-square = 2.17, df = 3, P = 0.538.*

#### Marital status

| Category | n | Gap events | Gap rate | 95% CI |
| --- | --- | --- | --- | --- |
| Divorced | 169 | 17 | 10.1% | 6.4%, 15.5% |
| Married | 783 | 92 | 11.7% | 9.7%, 14.2% |
| Single | 999 | 94 | 9.4% | 7.8%, 11.4% |
| Widowed | 195 | 23 | 11.8% | 8.0%, 17.1% |

*Omnibus chi-square test of independence: chi-square = 2.94, df = 3, P = 0.401.*

### eTable 2. Sensitivity analyses of the primary gap-rate estimate

*The primary estimate uses a 1.5-times-expected-interval gap threshold and the full analysis dataset (n = 2,469 transition-by-drug observations). Each row varies one analytic choice while holding the others at their primary-analysis value. The 95% CI for the primary row is the admission-cluster bootstrap interval, the primary confidence-interval method for the headline estimate; 95% CIs for all other rows are Wilson score intervals, the method used throughout for these secondary and sensitivity analyses (eMethods).*

| Analysis | n | Gap rate | 95% CI |
| --- | --- | --- | --- |
| Primary (1.5x threshold, full dataset) | 2,469 | 10.2% | 8.7%, 11.7% |
| 2.0x threshold | 2,469 | 7.1% | 6.1%, 8.2% |
| 3.0x threshold | 2,469 | 5.5% | 4.7%, 6.5% |
| One transition per admission (clustering sensitivity) | 1,583 | 11.1% | 9.7%, 12.8% |
| Switch-excluded (same-day medication change excluded; n excluded = 1,373) | 1,096 | 10.8% | 9.1%, 12.7% |
| Ward-only (excludes step-down/intermediate destinations) | 2,250 | 10.8% | 9.6%, 12.1% |

*IV-availability class comparison (chi-square test of independence): chi-square = 0.79, Cramer V = 0.018, P = 0.374.*

### eTable 3. Matched non-transfer control-interval comparison, full contingency detail

*Each transfer interval was paired with a non-transfer control interval from the same patient and drug and independently flagged as a gap using the same 1.5-times-expected-interval threshold (eMethods). The pre-transfer control interval is the interval between the second-to-last and last ICU-charted dose; the post-transfer control interval is the interval between the first and second floor-charted dose. Discordant and concordant counts refer to the paired gap flag on the transfer interval versus its control interval; McNemar P is from an exact, two-sided McNemar test on the discordant pairs; the rate-difference CI, not the McNemar P value, is the primary clustering-aware uncertainty estimate. Rate difference is the transfer-interval gap rate minus the control-interval gap rate; its 95% CI is from a 5,000-resample bootstrap clustered by hospital admission identifier (primary) or by patient identifier (sensitivity), restricted to this control-evaluable subset (eMethods).*

| Field | Pre-transfer control | Post-transfer control |
| --- | --- | --- |
| n | 2,247 | 2,356 |
| Transfer-interval gap rate | 7.5% | 8.9% |
| Control-interval gap rate | 1.7% | 2.5% |
| Discordant, gap across transfer only | 163 | 197 |
| Discordant, gap during control only | 33 | 46 |
| Concordant, gap in both | 6 | 13 |
| Concordant, gap in neither | 2,045 | 2,100 |
| McNemar P (exact, two-sided) | 7.26 × 10^-22^ | 1.92 × 10^-23^ |
| Rate difference, percentage points | 5.8 | 6.4 |
| 95% CI (admission-cluster bootstrap) | 4.4, 7.1 | 4.9, 7.9 |
| 95% CI (patient-cluster bootstrap, sensitivity) | 4.4, 7.2 | 4.8, 8.1 |

#### Control-evaluable-subset selection check

*The pre-transfer and post-transfer comparisons above are each restricted to the subset of the 2,469-observation analysis dataset with a constructible control interval of that type, and this subset is not a random sample of it (eMethods). This sub-table compares the gap rate among observations with a constructible control interval (the subset entering the comparison above) against observations without one. For the post-transfer comparison, the mechanism is identifiable: a post-transfer control interval requires two floor-charted doses of the same drug, so a never-resumed gap (no floor-charted dose by definition) and some early-discontinuation gaps cannot generate one, weighting the post-transfer control-evaluable subset toward milder gaps; the comparison above therefore characterizes this control-evaluable subset rather than the full analysis dataset.*

| Subset | n | Gap events | Gap rate | 95% CI |
| --- | --- | --- | --- | --- |
| Pre-transfer, has constructible control interval | 2,247 | 169 | 7.5% | 6.5%, 8.7% |
| Pre-transfer, no constructible control interval | 222 | 82 | 36.9% | 30.9%, 43.5% |
| Post-transfer, has constructible control interval | 2,356 | 210 | 8.9% | 7.8%, 10.1% |
| Post-transfer, no constructible control interval | 113 | 41 | 36.3% | 28.0%, 45.5% |

#### Post-transfer comparison, schedule-unchanged subset (sensitivity check)

*Restricts the post-transfer comparison above to the subset for which the floor-active dosing order was confirmed unchanged from the ICU-departure order (2,286 of 2,300 checked observations; eMethods), addressing the possibility that a later floor order with a different dosing frequency makes the ICU-departure-derived expected interval an imprecise yardstick for this comparison specifically.*

| Analysis | n | Transfer-interval gap rate | Control-interval gap rate | Discordant, transfer only | Discordant, control only | McNemar P (exact, two-sided) |
| --- | --- | --- | --- | --- | --- | --- |
| Post-transfer, full evaluable subset | 2,356 | 8.9% | 2.5% | 197 | 46 | 1.92 × 10^-23^ |
| Post-transfer, schedule-unchanged subset | 2,286 | 8.7% | 2.4% | 190 | 44 | 8.27 × 10^-23^ |

#### Threshold sensitivity (2.0x and 3.0x expected-interval thresholds)

*The pre-transfer and post-transfer matched comparisons above use the primary 1.5-times-expected-interval gap threshold; this sub-table repeats both comparisons at the alternative 2.0-times and 3.0-times thresholds used in the primary gap-rate sensitivity analysis (eTable 2), without altering which observations enter each comparison (eMethods).*

| Threshold | Comparison | n | Transfer rate | Control rate | Rate difference, pp | 95% CI (admission-cluster) | McNemar P |
| --- | --- | --- | --- | --- | --- | --- | --- |
| 2.0x | Pre-transfer | 2,247 | 4.6% | 1.0% | 3.6 | 2.6, 4.7 | 1.96 × 10^-14^ |
| 2.0x | Post-transfer | 2,356 | 5.8% | 1.0% | 4.8 | 3.6, 6.1 | 2.43 × 10^-21^ |
| 3.0x | Pre-transfer | 2,247 | 3.3% | 0.3% | 3.0 | 2.2, 4.0 | 1.50 × 10^-16^ |
| 3.0x | Post-transfer | 2,356 | 4.3% | 0.2% | 4.1 | 3.1, 5.3 | 1.38 × 10^-24^ |

#### Ward-only sensitivity (excludes step-down/intermediate destinations)

*The pre-transfer and post-transfer matched comparisons above include admissions transferred to step-down or intermediate-care destinations; this sub-table restricts both comparisons to the subset transferred to a regular medical/surgical floor only, addressing whether the matched-comparator excess is specific to standard floor destinations rather than an artifact of including step-down destinations (eTable 2).*

| Comparison | n | Transfer rate | Control rate | Rate difference, pp | 95% CI (admission-cluster) | McNemar P |
| --- | --- | --- | --- | --- | --- | --- |
| Pre-transfer | 2,034 | 7.9% | 1.9% | 6.0 | 4.6, 7.6 | 1.46 × 10^-20^ |
| Post-transfer | 2,141 | 9.6% | 2.7% | 6.9 | 5.3, 8.6 | 3.12 × 10^-23^ |

### eTable 4. Delay-duration distribution among administration gaps

*The primary summary is the excess delay beyond each gap's own expected dosing interval: the raw interdose interval, the interval in hours between the last confirmed ICU dose and the first confirmed floor dose, or, for a gap without a further confirmed dose, between the last confirmed ICU dose and hospital discharge, minus that observation's expected dosing interval (eMethods). The raw interdose interval itself is reported alongside as a secondary, absolute-elapsed-time reference. The lower panel reports the gap rate by the medication's expected dosing interval, with Wilson score 95% confidence intervals.*

#### Delay duration, by resumption status

**Excess delay beyond expected dosing interval (primary)**

| Gap category | n | Median, h | IQR, h | 90th percentile, h | Max, h |
| --- | --- | --- | --- | --- | --- |
| All gaps | 251 | 24.2 | 10.5, 58.7 | 131.0 | 710.5 |
| Resumed before discharge | 221 | 23.8 | 10.4, 51.2 | 118.8 | 710.5 |
| Not resumed before discharge | 30 | 51.9 | 21.4, 119.3 | 173.7 | 465.4 |

**Raw interdose interval (secondary, absolute-elapsed-time reference)**

| Gap category | n | Median, h | IQR, h | 90th percentile, h | Max, h |
| --- | --- | --- | --- | --- | --- |
| All gaps | 251 | 37.2 | 22.6, 71.6 | 143.0 | 722.5 |
| Resumed before discharge | 221 | 36.4 | 22.4, 64.5 | 131.9 | 722.5 |
| Not resumed before discharge | 30 | 61.9 | 33.4, 130.3 | 185.7 | 477.4 |

#### Gap rate by expected dosing interval

| Dosing interval | n | Gap events | Gap rate | 95% CI |
| --- | --- | --- | --- | --- |
| Every 6 hours | 42 | 7 | 16.7% | 8.3%, 30.6% |
| Every 8 hours | 201 | 56 | 27.9% | 22.1%, 34.4% |
| Every 12 hours | 1,844 | 158 | 8.6% | 7.4%, 9.9% |
| Every 24 hours | 382 | 30 | 7.9% | 5.6%, 11.0% |

### eTable 5. Antiseizure medication administration gap rate by medication

*Gap rate and Wilson score 95% confidence interval for each of the ten antiseizure medications in the analysis dataset (n = 2,469), sorted from lowest to highest gap rate, matching the ordering in Figure 2b of the main text.*

| Medication | n | Gap events | Gap rate | 95% CI |
| --- | --- | --- | --- | --- |
| Lacosamide | 256 | 13 | 5.1% | 3.0%, 8.5% |
| Topiramate | 67 | 5 | 7.5% | 3.2%, 16.3% |
| Levetiracetam | 1,274 | 106 | 8.3% | 6.9%, 10.0% |
| Lamotrigine | 190 | 17 | 8.9% | 5.7%, 13.9% |
| Zonisamide | 65 | 7 | 10.8% | 5.3%, 20.6% |
| Oxcarbazepine | 100 | 14 | 14.0% | 8.5%, 22.1% |
| Phenobarbital | 61 | 9 | 14.8% | 8.0%, 25.7% |
| Valproate | 165 | 27 | 16.4% | 11.5%, 22.8% |
| Phenytoin | 229 | 41 | 17.9% | 13.5%, 23.4% |
| Carbamazepine | 62 | 12 | 19.4% | 11.4%, 30.9% |
